# What (if anything) is going on? Examining longitudinal associations between social media use and mental ill-health among young people

**DOI:** 10.1101/2022.03.31.22273198

**Authors:** Yvonne Kelly, Baowen Xue, Cara Booker, Amanda Sacker, Rebecca Lacey, George Ploubidis, Praveetha Patalay

**Affiliations:** Research Department of Epidemiology and Public Health, University College London, 1-19 Torrington Place, London, WC1E 7HB, United Kingdom; Institute for Social and Economic Research (ISER), University of Essex, Wivenhoe Park, Colchester, Essex, CO4 3SQ UK; UCL Institute of Education, University College London, 20 Bedford Way, London, WC1H 0AL; Centre for Longitudinal Studies and MRC Unit for Lifelong Health and Ageing, University College London

## Abstract

**Objectives:** In mid-adolescence, to 1) examine cyclical associations between social media use and mental ill health by investigating longitudinal and bidirectional associations, dose response relationships, and changes in social media use and in mental health; 2) assess potential interaction effects between social media use and mental health with pre-existing early life vulnerabilities.

**Methods:** Longitudinal data on 12,114 participants from the Millennium Cohort Study on social media use, depressive symptoms, self-harm and early life risk factors were used.

**Results:** We found little support for the existence of cyclical relationships between social media use and mental health. Where detected, effect sizes were small. Dose response associations were seen in the direction of mental health to social media (depressive symptoms 1 time OR=1.22, 2 times OR=1.71; self-harm 1 time OR=1.17, 2 times OR=1.53), but not for social media to mental health. Changes in social media use and changes in mental health were not associated with each other. We found no evidence to suggest that either social media use or mental health interacted with pre-existing risk for mental ill health.

**Conclusions:** Findings highlight the possibility that observed longitudinal associations between social media use and mental health might reflect other risk processes or vulnerabilities and/or the precision of measures used. More detailed and frequently collected prospective data about online experiences, including those from social media platforms themselves will help to provide a clearer picture of the relationship between social media use and mental health.

## Introduction

Young people’s mental ill-health is a growing public health challenge ^1-4^ with deteriorations seen over the last decade, and it is likely that a range of factors underly these trends. Higher rates of mental ill health such as depression and anxiety, have coincided with the increased uptake of certain types of digital technology use, for example social media. There has been much scientific, as well as public interest in the potential for a causal role between social media use and mental ill-health particularly among young people. Overall, findings point to, at most, modest associations between the amount of time spent on social media platforms and mental ill-health,^5-9^ and where gender differences have been examined findings suggest associations are more apparent for girls, especially in early adolescence.^5,6,10,11^

However, there are clear limitations in the research undertaken to date, and important questions remain about i) whether these associations reflect causal effects, ii) the magnitude and direction of associations, and iii) whether there are vulnerable young people at particular risk of experiencing mental ill-health and potentially problematic social media use. To address these questions, studies need to tackle key limitations of extant research. Firstly, most findings are cross sectional and have shown between-group comparisons,^6,8^ thus preventing examination of potentially cyclical associations – it is possible that poor mental health in the first instance leads to higher social media use which in turn may lead to more mental health problems.^9,11-14^ Secondly, findings mostly use measures of time spent on social media platforms which in itself tells us little about individual experiences. For instance, if time spent on these platforms promotes connectedness and social support then it is unlikely to impact negatively on mental health. Conversely, experiences such as upward social comparisons, fear of missing out and negative interactions are most likely to be tied to poor mental health.^15,16^ Thirdly, few studies integrate measures of underlying risk for mental health problems such as structural disadvantage and adverse psychosocial circumstances^17-20^ and it may be that there are interplays between aspects and experiences of online environments with underlying off-line vulnerabilities that exacerbate or mitigate extant risk.^21^ In this paper we address some of the key limitations that have plagued prior studies by: a) looking longitudinally at social media use and mental health within individuals; b) using measures of different types of experiences on social media in addition to time spent on platforms e.g. compulsive use, and feeling happier and more connected than in real life; and c) investigating whether there is interplay between online experiences and pre-existing off-line vulnerability for mental ill-health and potentially problematic social media use.

Given the size of the public health challenge posed by mental ill health among young people, coupled with lifelong implications^22^, identifying potentially modifiable risk factors has the potential for substantial societal benefits across the lifecourse. If online experiences including those encountered on social media platforms are found to play a causal role, even a modest one, then it will be critical in developing preventive strategies.

This paper has two broad aims, the first is to tease apart the potential cyclical nature of associations between social media use and mental ill health (measured using depressive symptoms and self-harm) in mid-adolescence. We do this in three ways, by: i) assessing longitudinal and bi-directional pathways between social media use and mental ill-health in early and later adolescence; ii) investigating whether there are cumulative associations at play in either or both directions i.e. looking at whether ‘heavy’ social media use throughout adolescence is associated with mental ill-health in later adolescence, and vice versa; and iii) examining whether changes in social media use are associated with changes in markers of mental ill-health, and vice-versa. Our second aim is to assess potential interaction effects between social media use and experiences, and the impact of a range of pre-existing risk factors for adolescent mental ill-health. Here we consider the social determinants of young people’s mental health, including structural factors - socioeconomic disadvantage,^20^ and special educational needs and disability;^19^ and proximal psychosocial factors - parental mental health,^17^ physical punishment,^23^ and bullying.^18^ We examine these associations in both directions, by investigating whether associations between different aspects of social media use (lengthy hours, compulsiveness, feeling happier and more connected than in real life) with mental health interact with pre-existing vulnerability factors.

To do this we make use of data from the UK Millennium Cohort Study (MCS), a longitudinal nationally representative cohort. For all analyses we exploit the longitudinal design of the MCS by making use of multiple data sweeps. Our research questions (as depicted in Figure 1) are as follows:

**Figure 1.**
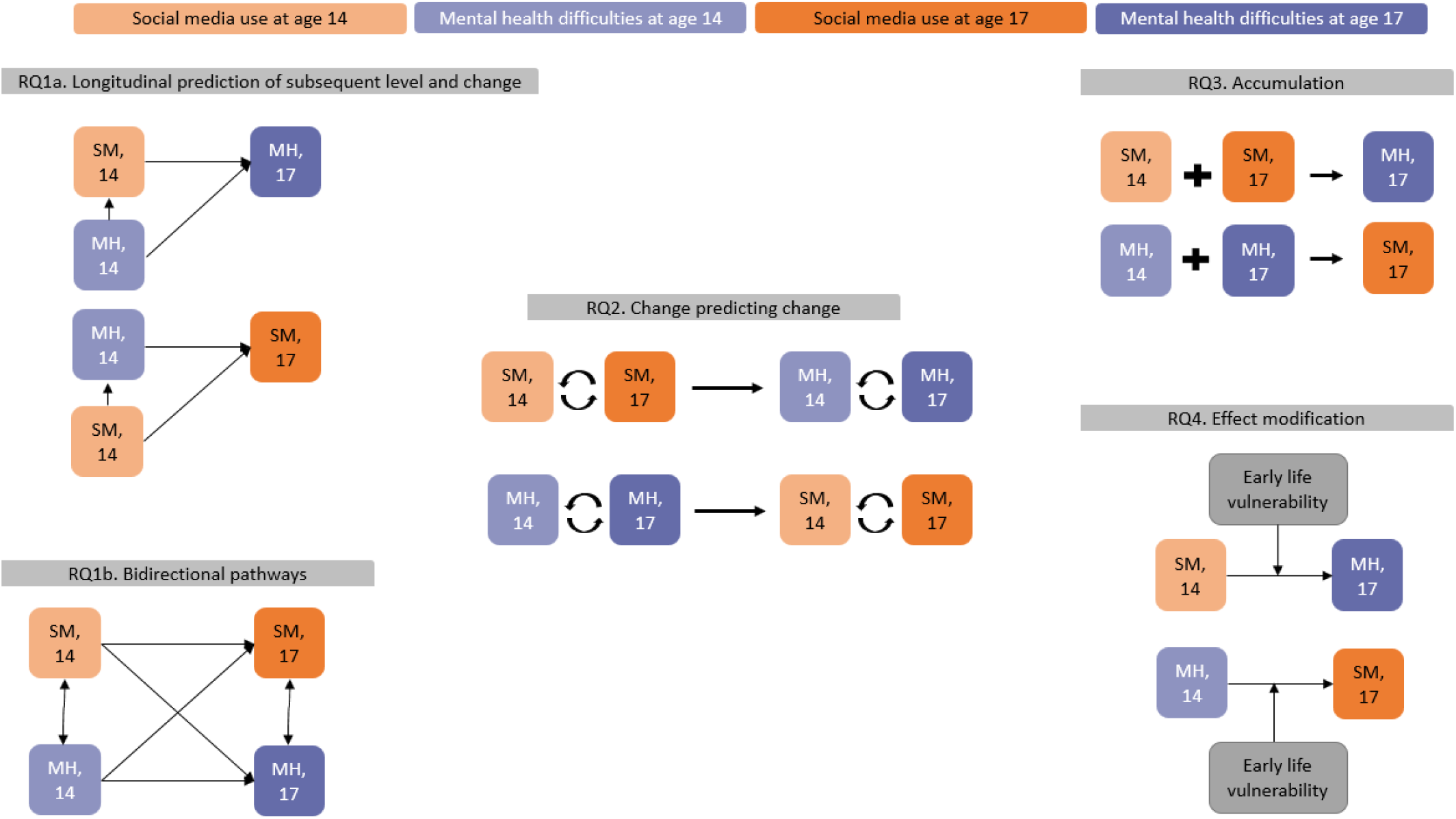
Schematic overview of the RQs and the models estimated to answer each.

RQ1 Are there (a) longitudinal and (b) bi-directional pathways between social media use and mental ill-health in mid-adolescence from age 14 to age 17?
RQ2 Are there cumulative associations between social media use and mental ill-health in either or both directions?
RQ3 Are changes in social media use between ages 14 and 17 associated with changes in markers of mental ill-health, and vice-versa?
RQ4 Do different aspects of social media use (lengthy hours, compulsiveness, feeling happier and more connected than in real life) interact with pre-existing vulnerability for mental ill-health, and vice-versa?

## Methods

### Millennium Cohort Study

The MCS is a UK nationally representative prospective cohort study of children born into 19244 families between September 2000 and January 2002 (http://www.cls.ioe.ac.uk/shared/get-file.ashx?id=1806&itemtype=document). Participating families in receipt of Child Benefit (98% of the population at the time of sampling) were selected from a random sample of electoral wards with a stratified sampling design to ensure adequate representation of all four UK countries, disadvantaged and ethnically diverse areas. The first sweep of data was collected when cohort members were around 9 months and the subsequent six sweeps of data were collected at ages 3, 5, 7, 11 14 and 17 years.

At ages 7, 11, 14 and 17 cohort members were interviewed, and at all data collection sweeps their carers were interviewed during home visits. Cohort members self-completed computer assisted questionnaires in private including items about social media use, mental health, and relationships with family and peers. Carers (the majority of whom were cohort member’s parents and for ease throughout are referred to as parents) answered questions about their socioeconomic circumstances, parenting strategies, and cohort member’s wellbeing throughout childhood including whether they had any disabilities and special educational needs. Interview data were available for 76.3% of families when cohort members were aged 14, and 73.6% of families when cohort members were aged 17.

### Mental ill-health

We include two indicators of mental ill-health: depressive symptoms and self-harm

#### Depressive symptoms

At age 14 participants completed the Short Mood and Feelings Questionnaire – short version (SMFQ) from which a summed score was created. The SMFQ comprises 13 items on affective symptoms in the last 2 weeks (see Box 1). At age 17 participants completed the Kessler – 6 items (see Box 1) from which a sum score was created. While the two measures predominantly capture symptoms of depression, there are some differences in the numbers and types of symptoms included with the 13-item SMFQ covering more symptoms by virtue of its length. Five items on each measure capture similar symptoms (hopelessness, restless/fidgety, worthlessness, depressed/miserable, too much effort). Sensitivity analyses conducted with these common symptoms yielded similar results to those using full measures. We use continuous depressive symptom measures - z scores (DS z), and a binary indicator of depressive symptoms with a cut point of 1 standard deviation above the mean to indicate a ‘high’ level of symptoms at both ages. In addition, sensitivity analyses using binary depressive symptom measures based on ‘caseness’ cut points from the original measures (≥ 12 for SMFQ, and ≥ 13 for Kessler – 6) yielded similar results to those using +1SD measures.

##### Box 1

Questionnaire items

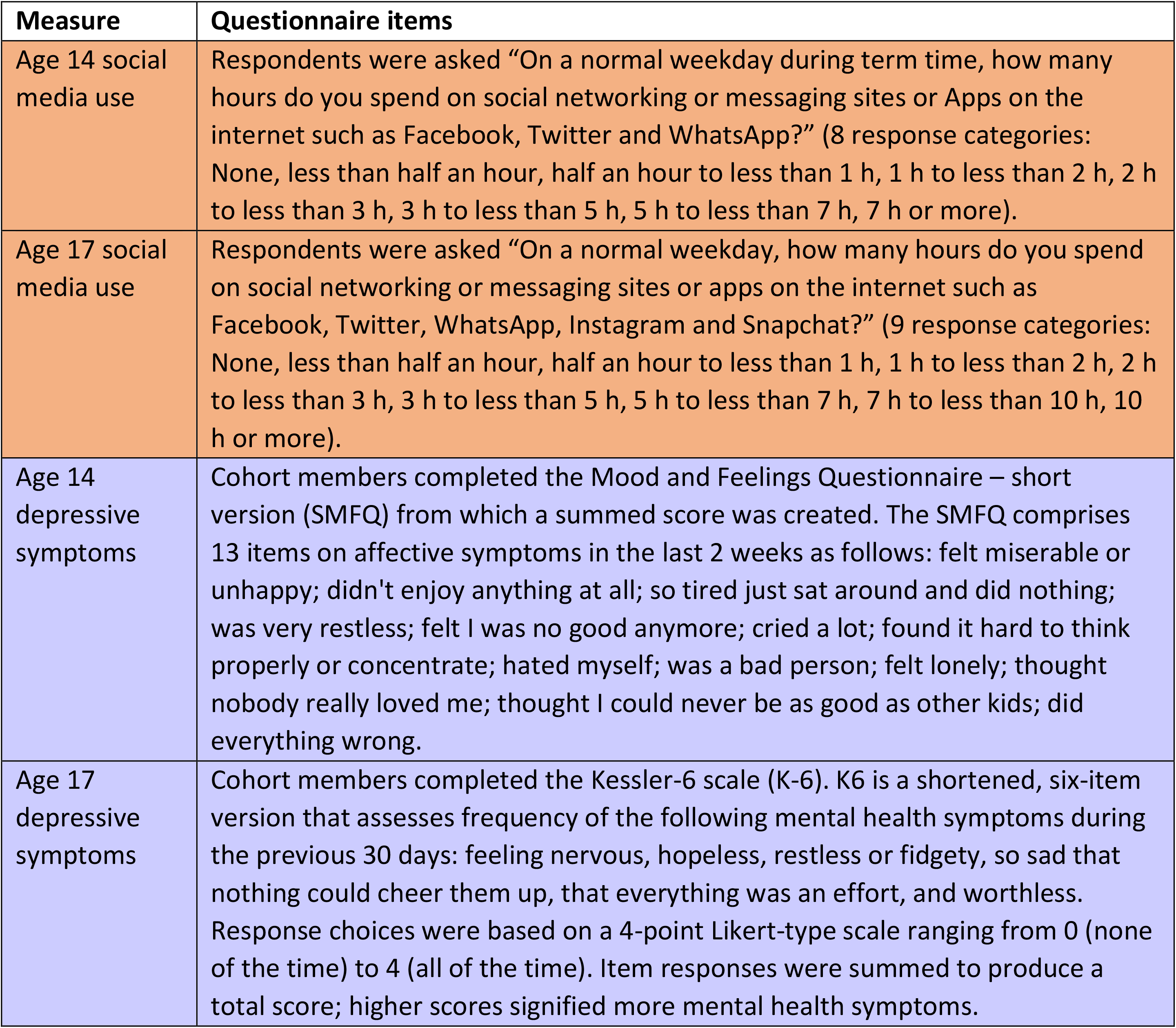

#### Self-harm

At age 14 participants were asked “In the past year have you hurt yourself on purpose in any way?” (yes vs no), and at age 17 participants were asked “During the last year, have you hurt yourself on purpose in any of the following ways?: Cut or stabbed yourself; Burned yourself; Bruised or pinched yourself; Taken an overdose of tablets; Pulled out your hair; Hurt yourself some other way” – answering yes to any of these was coded in a summary variable as reported self-harm.

#### Social media (SM) use

At ages 14 and 17 respondents were asked about their average hours of social media use on a weekday (for details of response categories see Box 1). The item used was developed by the Millennium Cohort Study team and is similar to items used in other large surveys including the UK Household Longitudinal Study and Ofcom. We use a continuous measure where possible to conserve statistical power, and a binary indicator to denote heavy SM use of ≥5 hours for some analyses. At age 17 respondents were asked an additional two questions capturing different aspects of social media experience – compulsiveness and connectedness– using, to what extent do you agree with the following statements? “I think I am addicted to social media” and “I’m more connected and happier online than I am in real life”, respectively. Responses were on a four point scale strongly agree through strongly disagree which we collapse to create binary variables, agree vs disagree.

#### Early lifecourse vulnerabilities

We generate variables to capture structural and psychosocial risk for poor adolescent mental health. Socioeconomic disadvantage was measured by the number of times cohort member families had an income below the poverty threshold (60% of median household income) between sweep 1 (9 months) and sweep 6 (age 14), collapsed to create a categorical variable (0/1-3/4-6 times). Special educational needs and disabilities (SEND) was measured from items asked from ages 5 through 14 years (‘gets help (in school) due to disability/problem’ and ‘has school told you your child has special needs’) to generate a variable - ever vs never. Parental mental health was measured using an average of parental Kessler-6 scores between age 3 and age 14. In the event one parent was not present in the household, only the other parent’s score was averaged. Physical punishment was measured using ‘How often smacks child when naughty’ between age 3 and 7 years, collapsed to create a binary variable (ever vs never). Experienced bullying by peers and siblings was measured using questions about frequency at ages 11 and 14 years. We use separate variables for peer (ever vs never) and sibling (ever, never, no siblings).

#### Covariates

We adjust for confounding variables in line with prior research^5,6^: age, family structure (one vs two parent) at age 14, emotional symptoms at age 11 (in last 4 weeks worried; sad; afraid or scared – items were used to create a summed score), in addition to the early lifecourse risk factors already described – socioeconomic disadvantage, SEND, parental mental health, and physical punishment. We did not adjust for experiences of bullying as in the case that causal processes are at play this has the potential to lie on the pathway between social media use and mental health.

#### Study sample and missing data

We analyse data on singleton-born cohort members for whom data on at least one marker of mental health or social media use are observed at either age 14 or age 17 (N=12,114). The largest percentage of missing data is the non-response of social media use at age 17 (43% missing), as these questions were collected in the young person online questionnaire (CAWI), completed after the home visit at age 17, which has a lower response rate than the face-to-face survey. The amount of missing data for each variable before imputation is shown in Supplementary Table S1.

To deal with missing data for all variables we use multiple imputation by chained equations, which account for uncertainty about missing values by imputing several values for each missing data point. We impute 50 data sets, and report consolidated results from all imputations using Rubin’s combination rules.^24^ Findings from sensitivity analyses with an analytic sample using listwise deletion are consistent with those using imputed data.

### Statistical analysis

Given the potential for gender differences in associations of interest we performed tests for interaction, but results indicate no differences thus we present gender adjusted findings for young women and men combined throughout. For all analyses, two indicators of mental health: depressive symptoms (depressive symptoms z scores, or a binary measure) and self-harm (binary) are analysed separately. All estimates are weighted with sample and non-response weights provided with the data.

#### RQ1 Are there (a) longitudinal and (b) bi-directional pathways between social media use and mental ill-health in mid adolescence from age 14 to age 17?

We first estimate a longitudinal regression model between social media use and mental health to test whether age 14 social media use is associated with mental health problems at age 17 (adjusting for confounders). We then further adjust for mental health at age 14 hence estimating whether age 14 social media use is associated with change in mental health between age 14 and age 17. Similar models are estimated for the opposite direction, that is whether mental health at age 14 is associated with social media use at age 17. Second, a cross-lagged panel model (path analysis) is used to investigate bi-directional associations between social media use and mental health. Cross-lagged models for depressive symptoms z score and self-harm are estimated using the generalised structural equation model (GSEM) command in Stata,^25,26^ which allows for continuous measures to be modelled using linear regression, and the use of logistic regression models for binary measures. We present models with adjustment for confounders (Model 1). Concurrent covariance/correlation cannot be used in a GSEM model, and thus two different models, with depressive symptoms z score or self-harm as the dependent variable (Model 2) and with social media hours as the dependent variable (Model 3), are shown.

#### RQ2 Are there cumulative associations between social media use and mental ill-health in either or both directions?

Using logistic regression models we test whether cumulative heavy social media use (5 hours or more per day) is associated with age 17 mental ill-health and whether cumulative mental ill-health is associated with age 17 heavy social media use. We generate three count variables for a) the number of times cohort members were heavy social media users, or b) had depressive symptoms, or c) self-harm (0/1/2 times).

#### RQ 3 Are changes in social media use between ages 14 and 17 associated with changes in mental ill-health, and vice-versa?

Here we estimate change in the dependent variable (DV) contingent on change in a particular independent variable (IV), i.e. ‘treatment’ effects, in the following four scenarios:

1. change in depressive symptoms z scores as the DV regressed on movement from non-heavy social media use at age 14 to heavy use at age 17 as the IV adjusted for age 14 social media hours.
2. onset of self-harm at age 17 as the DV regressed on change in social media hours adjusting for age 14 social media hours, and the sample for this model is restricted to those not reporting self-harm at age 14.

Looking at the other direction of influence

3. change in social media hours as the DV regressed on change in depressive symptoms score from age 14 to 17, adjusting for age 14 depressive symptoms score.
4. change in social media hours as the DV regressed on onset of self-harm at 17; as for scenario 2 the sample is restricted to those not reporting self-harm at age 14.

#### RQ4 Do different aspects of social media use interact with pre-existing vulnerability for poor mental health, and vice-versa?

First, we test age 17 cross-sectional associations between three aspects of social media use – hours, compulsiveness, feeling happier and more connected on social media than in real life – with mental health. Models first show the bivariate association between each aspect of social media use with mental health, and then mutually adjusted (with all 3 aspects entered together) associations are shown. Second, we test for interactions between social media use or mental health with markers of early life vulnerability by including an interaction term. We considered 6 early life vulnerability factors in combination with three social media exposures (hours, compulsiveness, more connected than in real life) and two mental health outcomes (depressive symptoms and self-harm) equalling 36 tests for interaction in each direction, 72 in total. To correct for false discoveries of multiple comparisons, we use the FDR correction^27^ with a corrected p<0.001 threshold.

## Results

Weekday hours spent on social media increased between ages 14 (mode=1-2 hours) and 17 (mode=3-5 hours) (Figure 2). Depressive symptoms and self-harm were associated with hours spent on social media. There were variations in the shape of these cross sectional associations by age, but overall, higher depressive symptom scores (Figure 3a) and higher prevalences of self-harm (Figure 3b) were observed among participants reporting more than 3 hours daily use.

**Figure 2.**
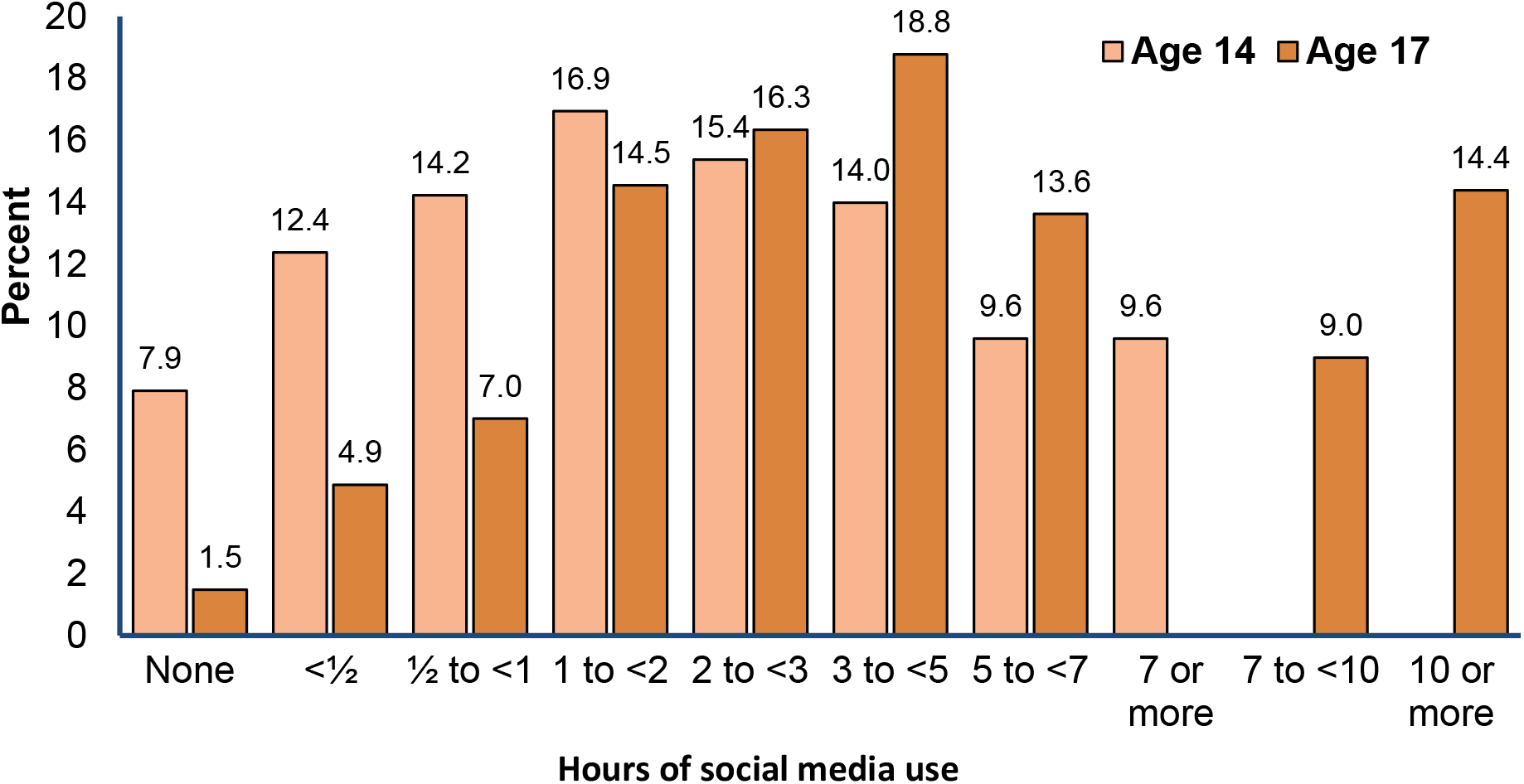
Weekday hours on social media at age 14 (censored at 7 hours) and age 17

**Figure 3a.**
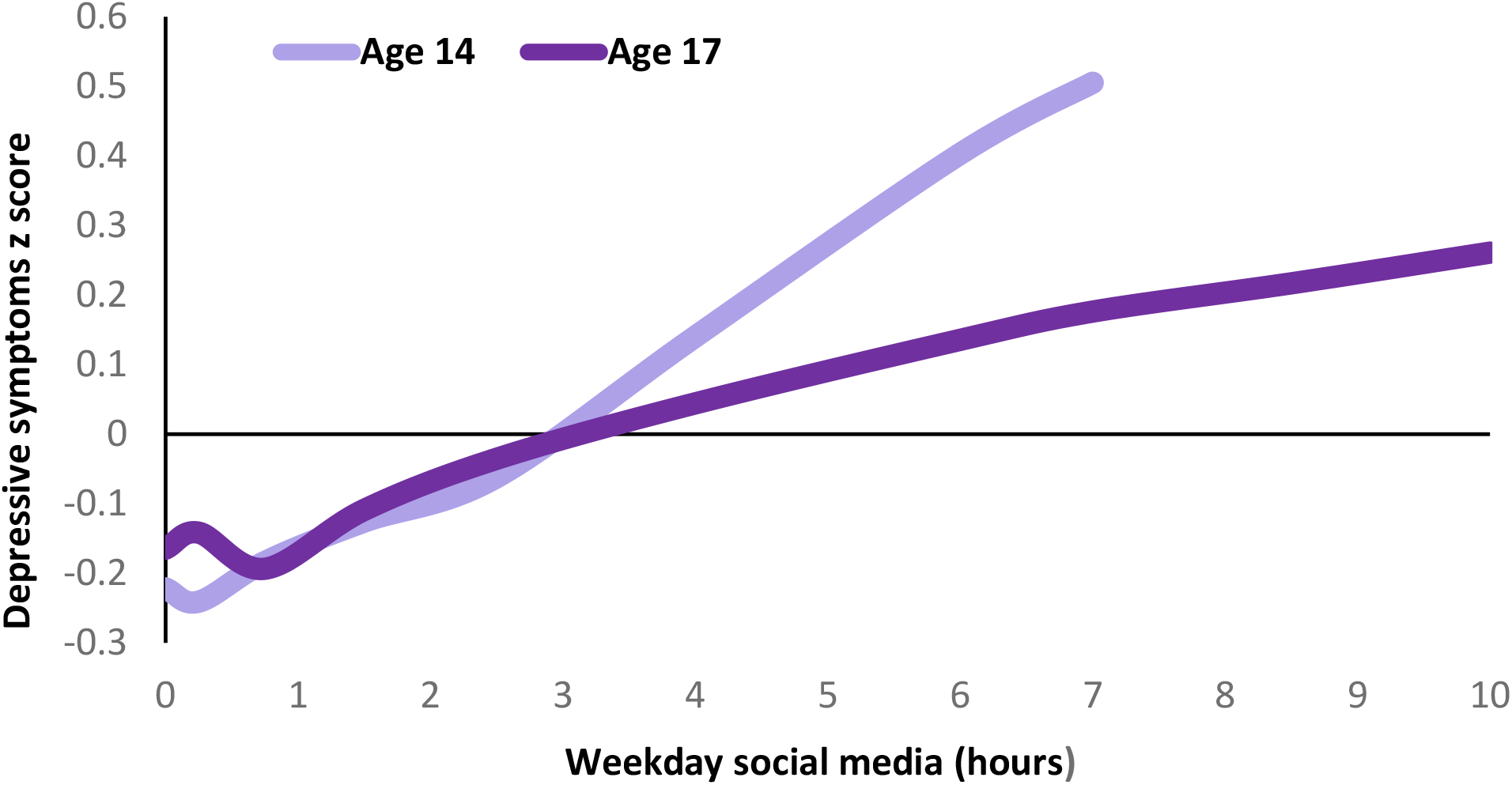
Social media hours and depressive symptoms (Age 14 social media use is censored at 7 hours)

**Figure 3b.**
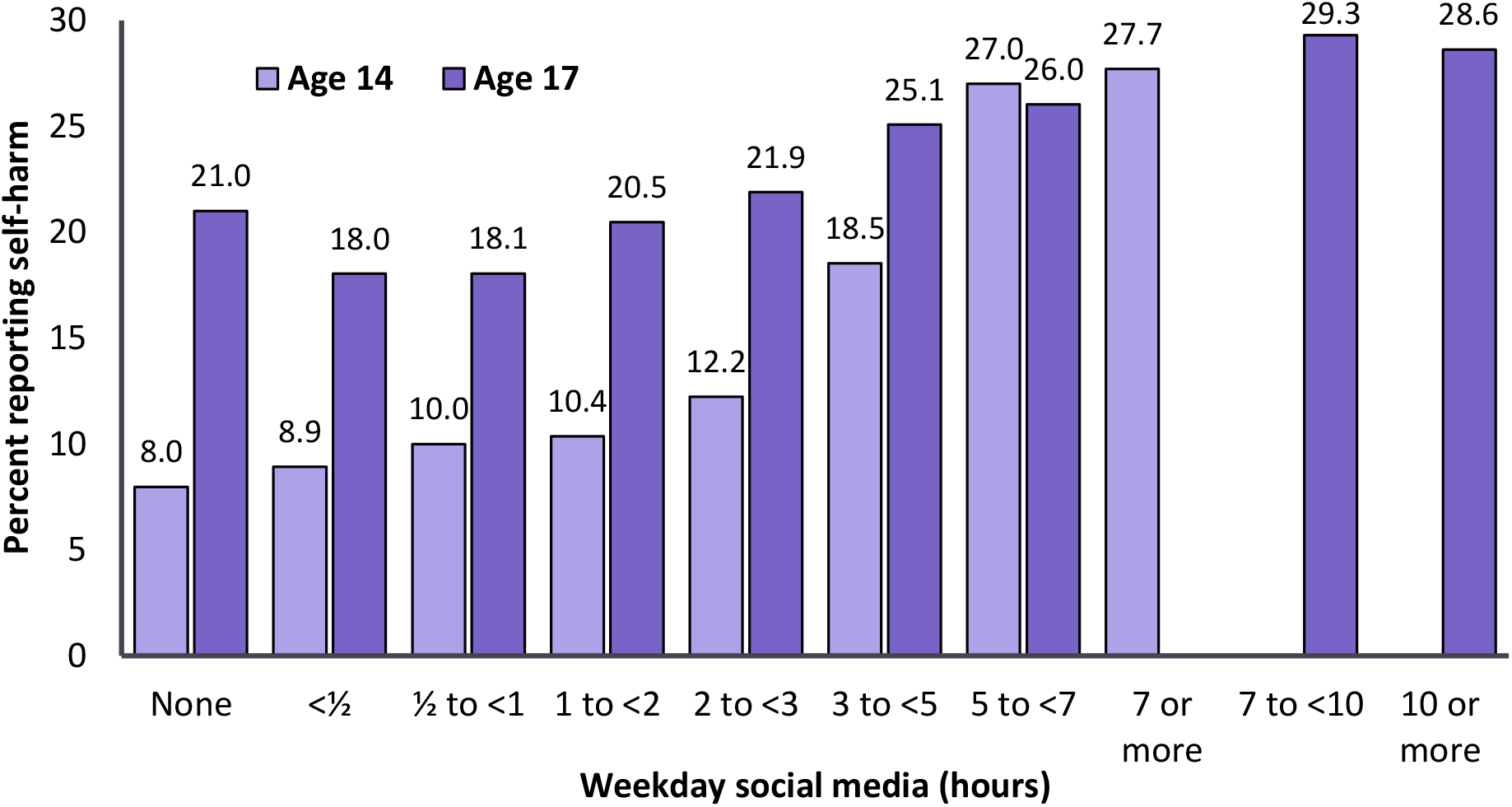
Social media hours and percent reporting self-harm (Age 14 social media use is censored at 7 hours)

### Are there longitudinal, and bi-directional pathways between social media use and mental ill-health from age 14 to age 17?

Social media hours at age 14 years were not associated with changes in either mental health outcome between ages 14 and 17 years, and similarly mental ill-health (whether depressive symptoms or self-harm) at age 14 was not associated with changes in social media use between ages 14 and 17 years (Table 1, Models 1 to 3). We did not observe associations in cross-lagged pathways between social media at 14 and mental health outcomes at age 17, and vice versa between mental health at 14 and social media use at age 17 (Figures 4 and 5 - single headed arrows represent directional associations/assumed causal relations).

**Table 1.**
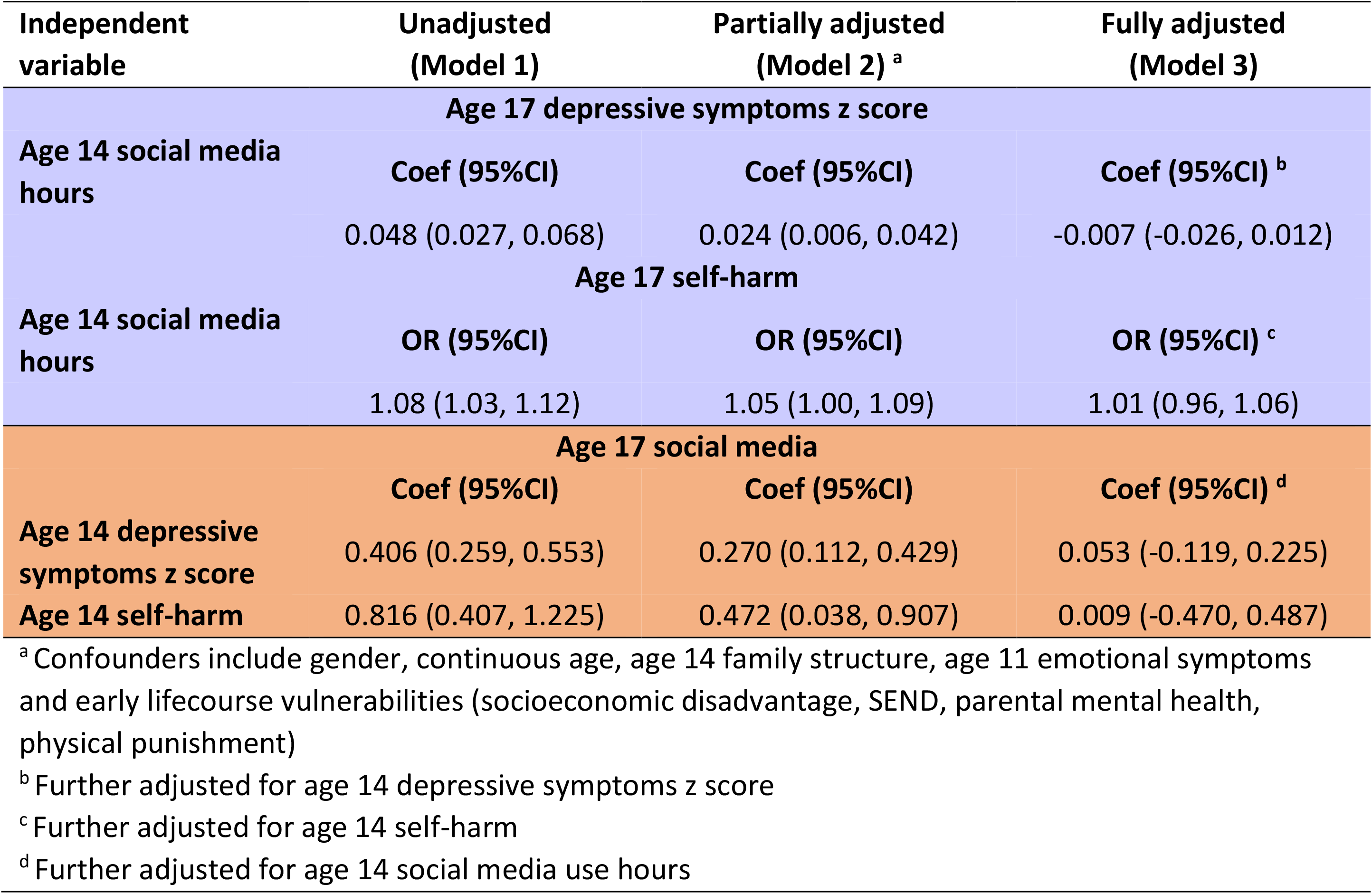
Longitudinal associations between social media use and mental health

**Figure 4.**
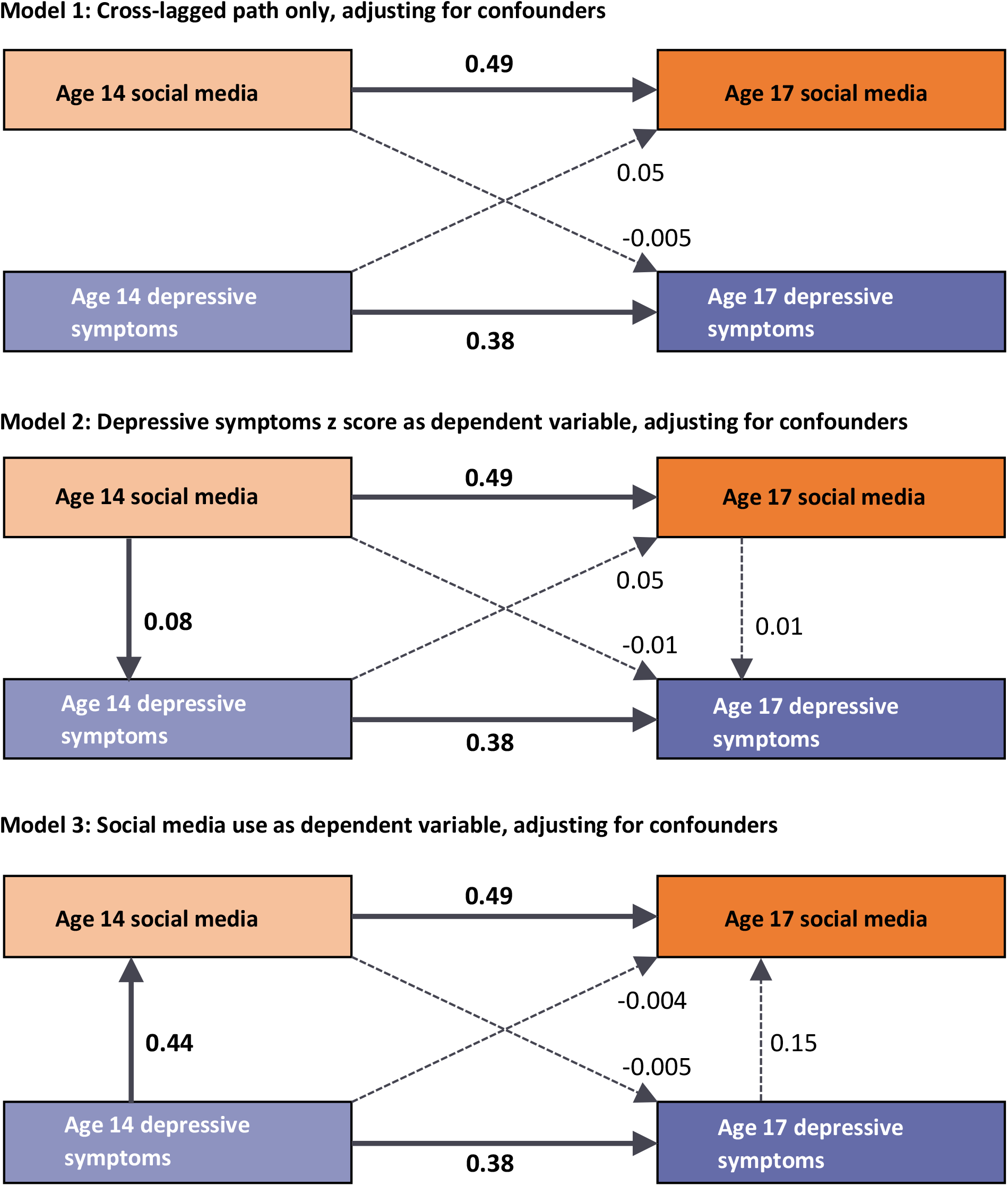
Bidirectional associations between social media use hours and depressive symptoms z score. Numbers next to the lines are coefficients. Coefficients with p<0.05 are shown in bold with solid lines, coefficients with p≥0.05 are shown with dashed lines. Adjusted confounders include gender, continuous age, age 14 family structure, age 11 emotional symptoms and early lifecourse vulnerabilities (socioeconomic disadvantage, SEND, parental mental health, physical punishment).

**Figure 5.**
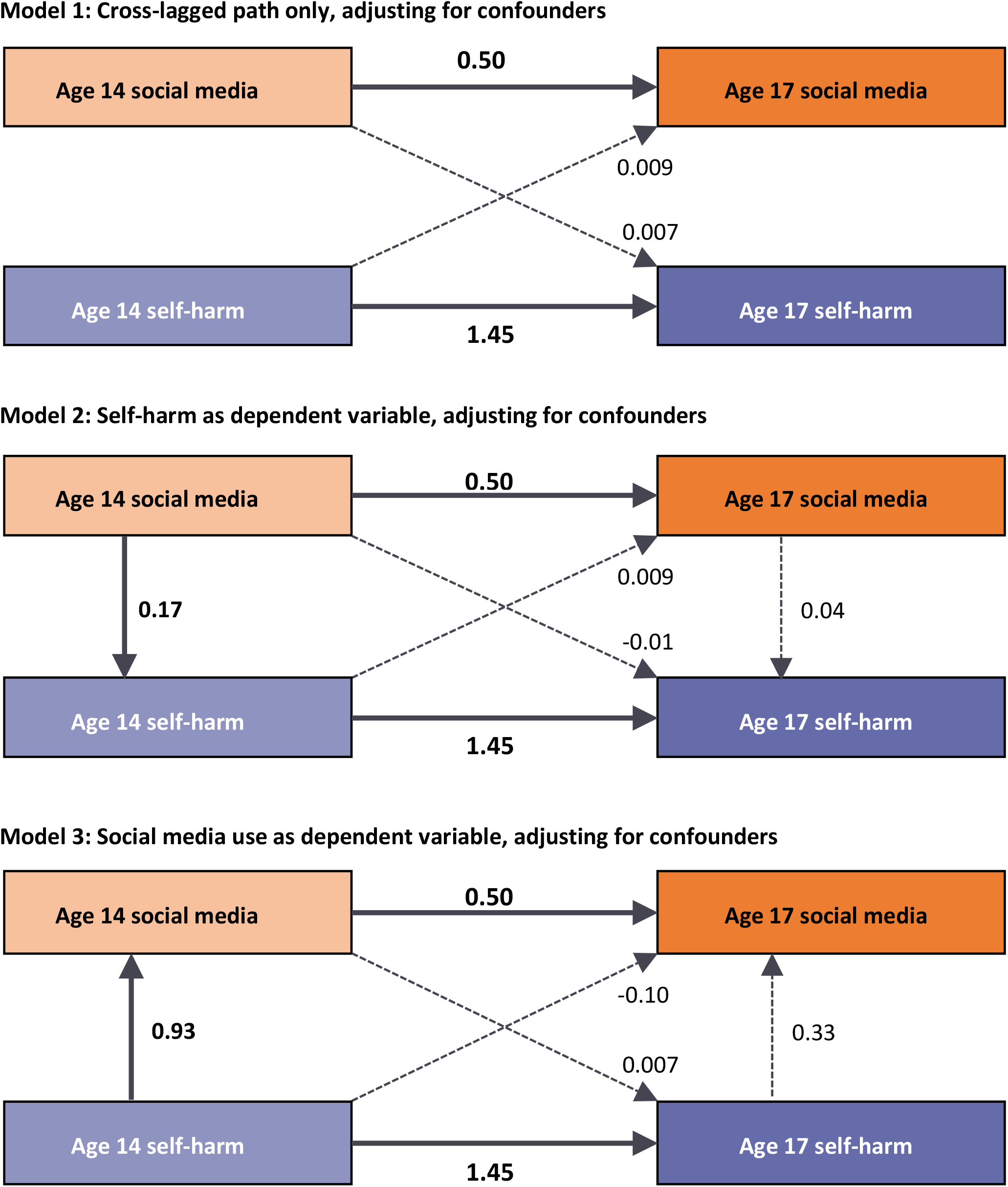
Bidirectional associations between social media use hours and self-harm. Numbers next to the lines are coefficients. Coefficients with p<0.05 are shown in bold with solid lines, coefficients with p≥0.05 are shown with dashed lines. Adjusted confounders include gender, continuous age, age 14 family structure, age 11 emotional symptoms and early lifecourse vulnerabilities (socioeconomic disadvantage, SEND, parental mental health, physical punishment).

### Are there cumulative associations between social media use and markers of mental health in either or both directions?

We observed weak support for dose-response associations in the direction of mental health to social media use though confidence intervals often overlap unity (Figure 6). For example, the odds of heavy social media use were as follows: depressive symptoms at 1 time OR=1.22, and at 2 times OR=1.71; self-harm at 1 time point OR=1.17, and at 2 time points OR=1.53.

**Figure 6.**
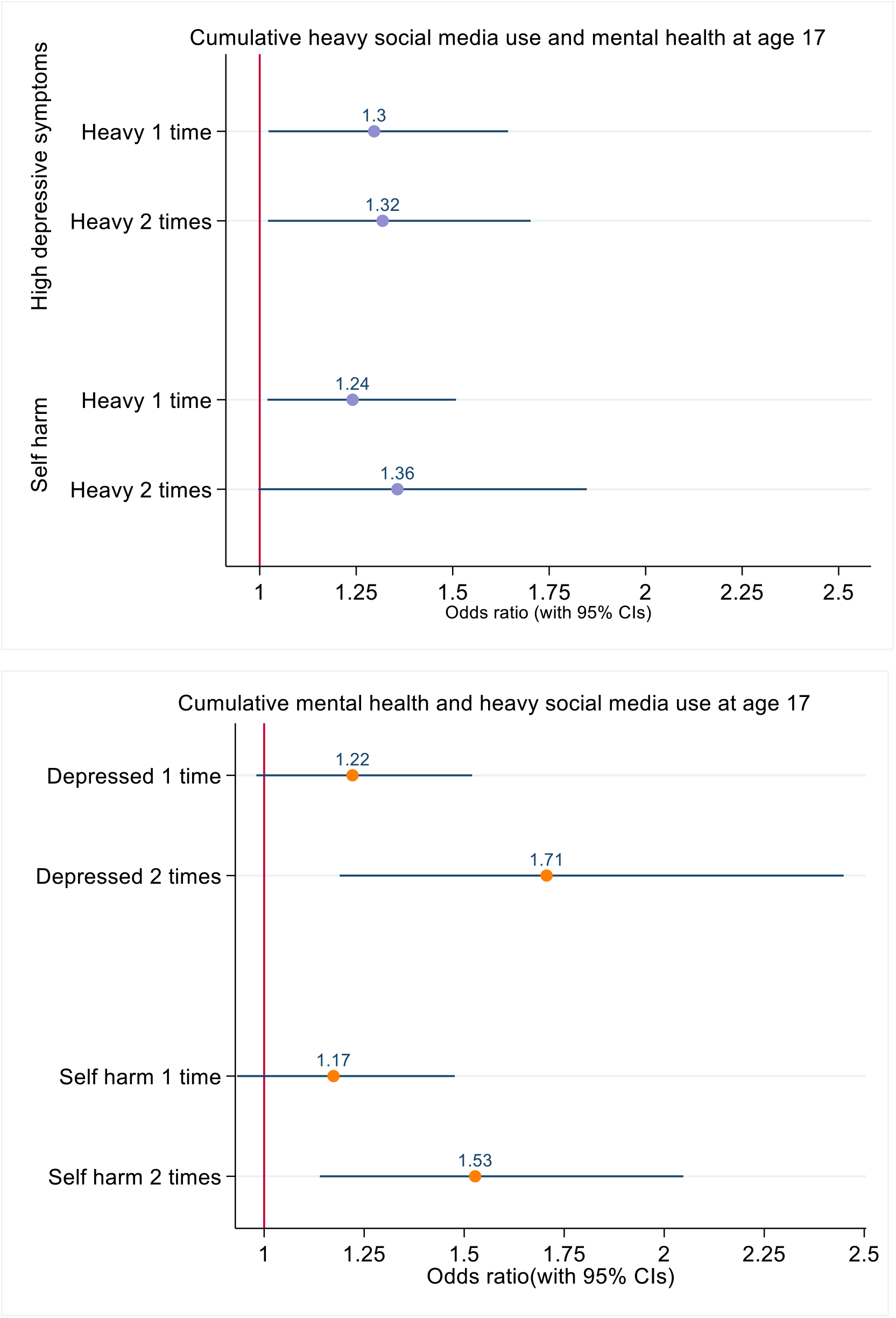
Cumulative associations between social media use and mental health. Adjusted confounders include gender, continuous age, age 14 family structure, age 11 emotional symptoms and early lifecourse vulnerabilities (socioeconomic disadvantage, SEND, parental mental health, physical punishment).

In the opposite direction, i.e. social media to mental health, there was no evidence of cumulative effects, the odds of depressive symptoms for heavy social media use were - at 1 time OR=1.30, and 2 times OR=1.32; and for self-harm at 1 time OR=1.24, and at 2 times OR=1.36.

### Are changes in social media use between age 14 and age 17 associated with changes in mental health and vice versa?

Overall, estimates were in the expected direction, although the magnitude of these associations were small and confidence intervals included the null (Table 2).

**Table 2.**
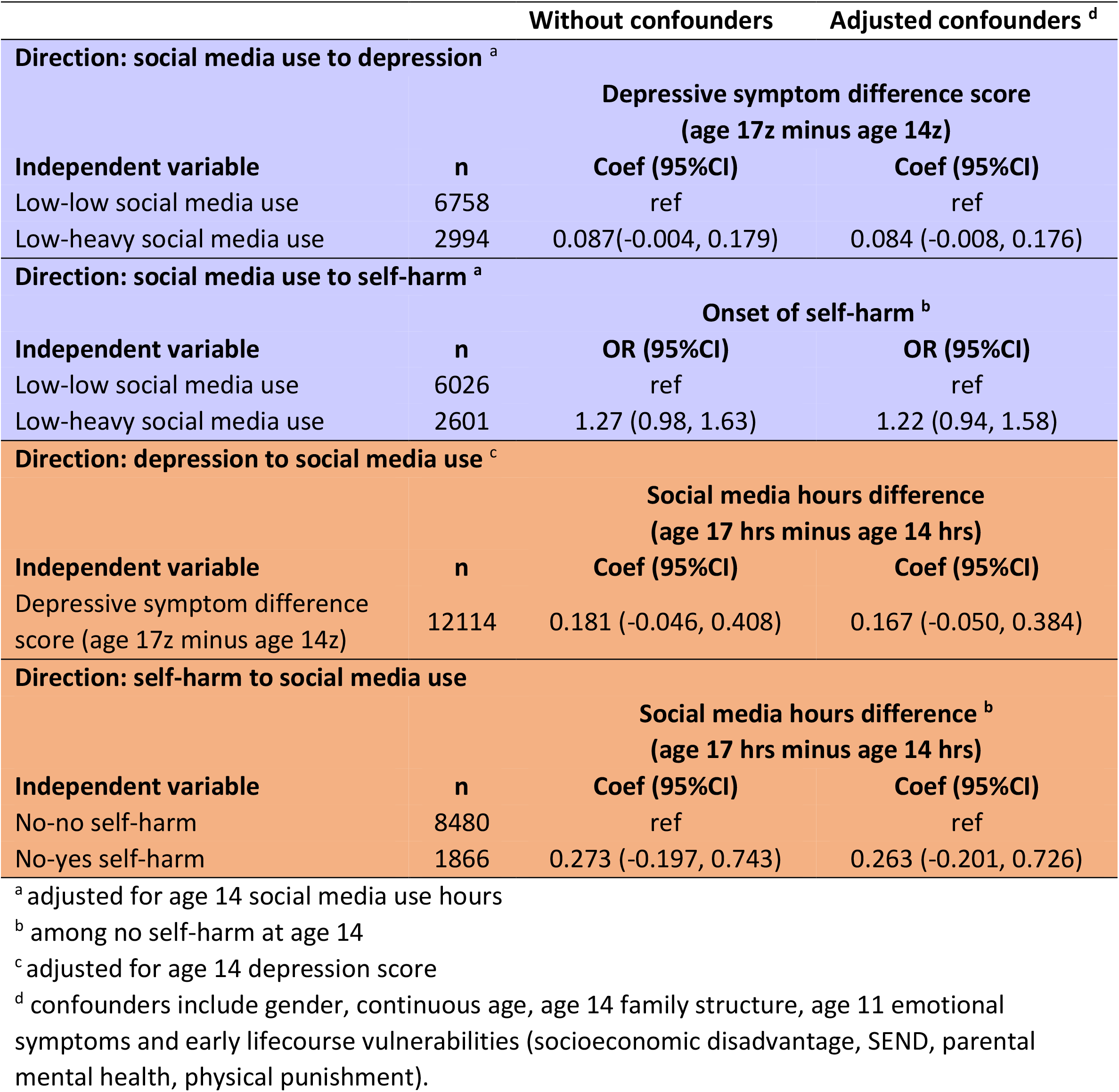
Associations between changes in social media use and changes in mental health

### Do different aspects of social media use interact with prior vulnerability for poor mental health, and vice-versa?

First, we tested cross-sectional associations between three aspects of social media use – hours of use, compulsiveness, feeling happier and more connected on social media than in real life with mental health outcomes at age 17 (Table 3). In mutually adjusted models feeling happier and more connected than in real life was strongly associated with both higher depressive symptom scores (coefficient=0.55) and self-harm (OR=2.15), but associations for compulsive use and hours of use were attenuated. We found little evidence of interactions when looking at associations between early life risk factors and mental health and these different aspects of social media use (Supplementary Tables S2-S3).

**Table 3.** Cross-sectional association between different aspects of social media use and depressive symptoms and self-harm at age 17

| Independent variable | Individually adjusted <sup>a</sup> | Mutually adjusted <sup>a b</sup> |
| --- | --- | --- |
| **Age 17 depressive symptoms z score** |  |  |
|  | **Coef (95%CI)** | **Coef (95%CI)** |
| Social media hours | 0.019 (0.005, 0.033) | 0.010 (-0.004, 0.022) |
| Social media compulsiveness | 0.121 (0.040, 0.202) | -0.014 (-0.089, 0.061) |
| Social media connectedness | 0.559 (0.471, 0.647) | 0.553 (0.466, 0.641) |
| **Age 17 self-harm** |  |  |
|  | **OR 95%CI** | **OR (95%CI)** |
| Social media hours | 1.04 (1.01, 1.08) | 1.03 (0.99, 1.06) |
| Social media compulsiveness | 1.25 (1.03, 1.50) | 0.99 (0.80, 1.21) |
| Social media connectedness | 2.20 (1.82, 2.66) | 2.15 (1.78, 2.60) |

<sup>a</sup> Confounders include gender, continuous age, age 17 family structure, age 11 emotional symptoms, physical punishment and early lifecourse vulnerabilities (socioeconomic disadvantage, SEND, parental mental health)
<sup>b</sup> Social Media Hours /Social Media Compulsiveness/Social Media Connectedness mutually adjusted

## Discussion

Among young people aged 14-17 years living in the UK, we found little support for the existence of cyclical relationships between social media use and mental health. Two of the three approaches used to test this hypothesis yielded estimates weakly supportive of a cyclical association, but effect sizes were small. Firstly, in longitudinal models, when we accounted for confounding variables, observed pathways were attenuated; secondly, there was a hint of dose response associations in the direction of mental health to social media, but not for social media to mental health; thirdly changes in social media use and changes in mental health were not consistently associated with each other. These findings highlight the very real possibility that there is no causal link between hours of social media use and mental ill-health in either direction. However, null findings might reflect the relevance of the measures available as length of time spent online might not matter so much, rather it is the nature of online encounters and experiences that are most likely to have an impact on young people’s mental health. Further here, in cross sectional analysis we found that young people who reported feeling happier and more connected online than in real life had higher levels of depressive symptoms and were more likely to report self-harm. On the other hand reported compulsive use was not associated with mental ill health when covariates were taken into account. Another potential explanation is due to the timeframe under study as there were three years between data collections and we might not expect to detect associations over this relatively lengthy gap in observations. We found no consistent evidence to suggest that either social media experiences or mental health in youth interacted with early life risk for mental ill health.

In keeping with other longitudinal studies, where we did detect associations, these were modest in size. ^5,7,9,11-14,28^ If a true association were to exist between social media use and mental health the potential underlying mechanisms are likely to be complex, and not necessarily related to the number of hours of use. On one hand, there are undoubtedly numerous benefits to be gained from using social media including making connections, social support and information acquisition. On the other hand, there could be downsides resulting from upward social comparisons, fear of missing out, negative interactions and encountering troubling content.^15,16^ Our observation that young people with sustained poor mental health are likely to spend more time using social media is consistent with studies showing that depressed youth seek help and support online.^29,30^ It has been proposed that associations between use of social media and mental health might be particular to sub-groups. Interestingly, we did not find evidence to support the hypothesis that social media use compounds effects of early life off-line risk.^21^ To test this hypothesis we examined a range of social determinants of mental health for young people, but there was no evidence to suggest that dimensions of social media use looked at in the present study interacted with off-line risk.

Our study has distinct strengths. Firstly, we used data from a large scale representative contemporary UK setting making our findings generalisable to the wider adolescent population in the UK. Secondly, we were able to examine associations longitudinally using multiple statistical methods, with different strengths and limitations, hence allowing us to present a multi-faceted and nuanced view of these associations - by investigating bidirectional associations (RQ1), considering dose response relationships (RQ2), and by looking at changes in both social media use and in mental health (RQ3). Thirdly, we were able to integrate data into our study not typically used on this research topic, for instance, on different aspects of social media use, including young people’s experiences, in addition to a range of early life risk factors for poor adolescent mental health (RQ4). To our knowledge, this is the first paper to have investigated the potential for interaction effects of social media with multiple aspects of early life vulnerability for poor adolescent mental health.

On the other hand, our study has distinct limitations, including that self-reported data of time spent on social media is crude and lacks accuracy,^31^ and use may be especially difficult to estimate in time categories as social media are not clearly delineated, unlike other forms of screen activities such as TV viewing and playing games. It may be that there is a risk of reporting bias for time spent on social media among young people experiencing mental ill health such that they under or over report time spent.^32^ However, the questions used were similar to those from other large-scale population based surveys including the UK Household Longitudinal Study^5^ and Ofcom^33^ and estimates of time spent using social media presented in our paper are consistent with those reported from other UK studies.^5,34^ We were able to go beyond self-reported time use to incorporate measures of experiences – compulsion and of feeling happier and more connected than in real life – however these measures were only available for a single time point preventing causal inference. Another limitation was that the measures of mental health used were different across time points thus potentially introducing measurement error. The relatively lengthy time lapse between data collections is also a limitation for the current study as it may be that shorter intervals between observations would have yielded different findings. Finally, as with most observational studies, there is the possibility that any positive findings could be a function of unmeasured confounding.

## Conclusions

There are many benefits to be gained for young people by engaging online but there may well be downsides too. The findings of our study highlight the possibility that any observed association between social media and mental health might reflect other risk processes or vulnerabilities. Thus, our findings, and those of others, highlight the likely complexity of mechanisms at play. Future research using more detailed and frequently harvested objectively collected prospective data including those from social media platforms themselves, along with more detailed accounts of online experiences, will help to provide a more comprehensive picture of the relationship between social media use and young people’s mental health. Given the short- and long-term implications of having poor mental health, improving our understanding of underlying processes could help identify opportunities for interventions with benefits across the lifecourse.

## Supporting information

Strobe checklist

## Data Availability

All data produced in the present study are available upon reasonable request to the authors

https://beta.ukdataservice.ac.uk/datacatalogue/series/series?id=2000031

## Acknowledgements

We would like to thank the Millennium Cohort Study families for their time and cooperation, as well as the Millennium Cohort Study team at the Institute of Education. The Millennium Cohort Study is funded by grants from Economic and Social Research Council.

## Ethical permission

Ethical approval was not required for this study as the analysis involved secondary analysis of publicly available data.

## Funding acknowledgments

During the conduct of the study YK and AS received funding from Economic and Social Research Council (ES/R008930/1). YK, PP and BZ received funding from Medical Research Council (MR/W002450/1). CB received funding from Economic and Social Research Council (RES-586-47-0001/RES-586-47-0002). RL received funding from Economic and Social Research Council (ESRC ES/P010229/1)

## Author contributions

YK, BX and PP designed the study. BX conducted data analysis. YK drafted the manuscript. BX, PP, AS, RL, CB and GP provided comments on drafts of the manuscript.

## Data access statement

Data are available on request from the authors.

**RQ1. What are the longitudinal and bidirectional pathways between social media use and mental health from age 14 to age 17?**

**RQ1 b Bidirectional associations between social media use and mental health**

**RQ2 Are there cumulative associations between social media use and markers of mental health in either or both directions?**

**RQ3 Are changes in social media use between age 14 & age 17 associated with changes in mental health and vice versa?**

## Supplementary materials

**Table S1.** Percentage of missing data before multiple imputation

|  | % missing (total n=12114) |
| --- | --- |
| **Age 11** |  |
| Emotional symptom | 11.29 |
| **Age 14** |  |
| Depressive symptoms | 9.91 |
| Self-harm | 9.09 |
| Social media use hours | 7.29 |
| Continuous age | 6.38 |
| Family structure | 6.39 |
| Socioeconomic disadvantage | 6.48 |
| Gender | 0 |
| **Age 17** |  |
| Depressive symptoms | 18.64 |
| Self-harm | 20.48 |
| Social media use hours | 43.23 |
| Social media compulsiveness | 43.26 |
| Social media connected | 43.38 |
| Continuous age | 16.79 |
| Family structure | 20.22 |
| Gender | 0 |
| **Early life vulnerabilities** |  |
| Socioeconomic disadvantage | range 3.89 – 9.38 across waves |
| SEND | range 7.32 – 8.03 across waves |
| Parental mental health | range 10.86 – 17.95 across waves |
| Physical punishment | range 13.92 – 20.06 across waves |
| Bullying by peers | range 6.52 – 8.8 across waves |
| Bullying by siblings | range 6.52 – 8.86 across waves |

**Table S2.** RQ4 Interactions - social media and markers of early life vulnerability on mental health as dependent variable

|  | Social media hours interactions |  | Social media compulsiveness interactions |  | Social media connectedness interactions |  |
| --- | --- | --- | --- | --- | --- | --- |
|  | Depressive symptoms z<br>p value | Self-harm<br>p value | Depressive symptoms z<br>p value | Self-harm<br>p value | Depressive symptoms z<br>p value | Self-harm<br>p value |
| **Socioeconomic disadvantage** |  |  |  |  |  |  |
| # 1-3 times | 0.421 | 0.817 | 0.554 | 0.538 | 0.423 | 0.898 |
| # 4-6 times | 0.865 | 0.521 | 0.989 | 0.278 | 0.777 | 0.233 |
| **SEND** | 0.432 | 0.694 | 0.980 | 0.503 | 0.901 | 0.874 |
| **Parental mental health** | 0.536 | 0.697 | 0.461 | 0.515 | 0.551 | 0.483 |
| **Physical punishment** | 0.761 | 0.771 | 0.886 | 0.647 | 0.313 | 0.440 |
| **Bullied by peers** | 0.546 | 0.160 | 0.982 | 0.225 | 0.411 | 0.563 |
| **Bullied by siblings** |  |  |  |  |  |  |
| # ever most days | 0.861 | 0.422 | 0.653 | 0.356 | 0.848 | 0.915 |
| # no sibling at ages 11&14 | 0.669 | 0.258 | 0.722 | 0.556 | 0.794 | 0.890 |

**Table S3.** RQ4 Interactions - mental health and markers of early life vulnerability on social media as dependent variable

|  | Depressive symptoms z score interactions |  |  | Self-harm interactions |  |  |
| --- | --- | --- | --- | --- | --- | --- |
|  | Heavy social media use<br>p value | Social media compulsiveness<br>p value | Social media connectedness<br>p value | Heavy social media use<br>p value | Social media compulsiveness<br>p value | Social media connectedness<br>p value |
| **Socioeconomic disadvantage** |  |  |  |  |  |  |
| # 1-3 times | 0.833 | 0.897 | 0.146 | 0.793 | 0.721 | 0.903 |
| # 4-6 times | 0.556 | 0.338 | 0.146 | 0.742 | 0.384 | 0.342 |
| **SEND** | 0.335 | 0.519 | 0.199 | 0.533 | 0.588 | 0.940 |
| **Parental distress score** | 0.529 | 0.785 | 0.795 | 0.430 | 0.533 | 0.408 |
| **Smack** | 0.668 | 0.694 | 0.514 | 0.782 | 0.843 | 0.435 |
| **Bully by peers** | 0.769 | 0.645 | 0.053 | 0.087 | 0.158 | 0.609 |
| **Bully by siblings** |  |  |  |  |  |  |
| # ever most days | 0.562 | 0.371 | 0.210 | 0.590 | 0.344 | 0.918 |
| # no sibling at ages 11&14 | 0.648 | 0.658 | 0.428 | 0.387 | 0.705 | 0.606 |

## References

1. Patton G, Borschmann R. Responding to the adolescent in distress. The Lancet.2017;390(10094):536–538.

2. Department of Health. Future in Mind: Promoting, protecting and improving our children and young people’s mental health and wellbeing. London: Department of Health;2015.

3. Centers for Disease Control and Prevention NCfIPaC. Webbased Injury Statistics Query and Reporting System (WISQARS). www.cdc.gov/injury/wisqars. Published 2018. Accessed 18 June 2020.

4. Mojtabai R, Olfson M, Han B. National Trends in the Prevalence and Treatment of Depression in Adolescents and Young Adults. Pediatrics. 2016;138(6):e20161878.

5. Booker CL, Kelly YJ, Sacker A. Gender differences in the associations between age trends of social media interaction and well-being among 10-15 year olds in the UK. BMC Public Health. 2018;18(1):321.

6. Kelly Y, Zilanawala A, Booker C, Sacker A. Social Media Use and Adolescent Mental Health: Findings From the UK Millennium Cohort Study. EClinicalMedicine. 2018;6:59–68.

7. Thorisdottir IESR.; Asgeirsdottir, B. B.; Allegrante, J. P.; Sigfusdottir, I.D. Active and Passive Social Media Use and Symptoms of Anxiety and Depressed Mood Among Icelandic Adolescents. Cyberpsychology, Behavior, and Social Networking. 2019;22(8):535–542.

8. Twenge JM, Joiner TE, Rogers ML, Martin GN. Increases in Depressive Symptoms, Suicide-Related Outcomes, and Suicide Rates Among U.S. Adolescents After 2010 and Links to Increased New Media Screen Time. Clinical Psychological Science. 2017;6(1):3–17.

9. Schemer C, Masur PK, Geiß S, Müller P, Schäfer S. The Impact of Internet and Social Media Use on Well-Being: A Longitudinal Analysis of Adolescents Across Nine Years. J Comput-Mediat Comm. 2020.

10. Barthorpe A, Winstone L, Mars B, Moran P. Is social media screen time really associated with poor adolescent mental health? A time use diary study. J Affect Disord. 2020;274:864–870.

11. Orben A, Dienlin T, Przybylski AK. Social media’s enduring effect on adolescent life satisfaction. Proc Natl Acad Sci U S A. 2019;116(21):10226–10228.

12. Oberst U, Wegmann E, Stodt B, Brand M, Chamarro A. Negative consequences from heavy social networking in adolescents: The mediating role of fear of missing out. J Adolesc. 2017;55:51–60.

13. Boers E, Afzali MH, Newton N, Conrod P. Association of Screen Time and Depression in Adolescence. JAMA Pediatrics. 2019;173(9):853–859.

14. Heffer T, Good M, Daly O, MacDonell E, Willoughby T. The Longitudinal Association Between Social-Media Use and Depressive Symptoms Among Adolescents and Young Adults: An Empirical Reply to Twenge et al. (2018). Clinical Psychological Science. 2019;7(3):462–470.

15. Clark JL, Algoe SB, Green MC. Social Network Sites and Well-Being: The Role of Social Connection. Current Directions in Psychological Science. 2018;27(1):32–37.

16. Seabrook EM, Kern ML, Rickard NS. Social Networking Sites, Depression, and Anxiety: A Systematic Review. JMIR Ment Health. 2016;3(4):e50.

17. Bevilacqua LKY.; Heilmann, A.; Priest, N.; Lacey, R. Adverse childhood experiences and trajectories of internalising, externalising, and prosocial behaviours from childhood to adolescence. J Am Acad Child Adolesc Psychiatry. In Press.

18. Patalay P. Changes in peer and sibling victimization in early adolescence: Longitudinal associations with multiple indices of mental health in a prospective birth cohort study. Submitted.

19. Office for National Statistics. Three years on: Survey of the development and emotional well-being of children and young people. 2008.

20. Reiss F. Socioeconomic inequalities and mental health problems in children and adolescents: a systematic review. Soc Sci Med. 2013;90:24–31.

21. Odgers CL, Jensen MR. Annual Research Review: Adolescent mental health in the digital age: facts, fears, and future directions. Journal of Child Psychology and Psychiatry. 2020;61(3):336–348.

22. Kessler RC, Amminger GP, Aguilar-Gaxiola S, Alonso J, Lee S, Üstün T. Age of onset of mental disorders: a review of recent literature. Curr Opin Psychiatry. 2007;20(4):359–364.

23. Heilmann AGL.; Kelly, Y.; van Turnhout, J.; Watt, R.; Mehay, A.;. Physical punishment narrative review. The Lancet. 2021.

24. Rubin DB. Multiple imputation for nonresponse in surveys. Vol 81. Hoboken, New Jersey: John Wiley & Sons; 2004.

25. Muthén B. A general structural equation model with dichotomous, ordered categorical, and continuous latent variable indicators. Psychometrika. 1984;49(1):115–132.

26. StataCorp. Structural Equation Modelling Reference Manual (Release 17). In: College Station, Texas: Stata Press; 2021: https://www.stata.com/manuals/sem.pdf.

27. Newson RB. Frequentist q-values for multiple-test procedures. Stata Journal. 2010;10(4):568–584.

28. Course-Choi J, Hammond L. Social Media Use and Adolescent Well-Being: A Narrative Review of Longitudinal Studies. Cyberpsychology, Behavior, and Social Networking. 2020.

29. Rideout VF, Susannah; and Well Being Trust, . Digital Health Practices, Social Media Use, and Mental Well-Being Among Teens and Young Adults in the U.S. 2018. https://digitalcommons.psjhealth.org/publications/1093.

30. Pretorius C, Chambers D, Coyle D. Young People’s Online Help-Seeking and Mental Health Difficulties: Systematic Narrative Review. J Med Internet Res. 2019;21(11):e13873.

31. Ellis DA, Davidson BI, Shaw H, Geyer K. Do smartphone usage scales predict behavior? International Journal of Human-Computer Studies. 2019;130:86–92.

32. Sewall CJR, Bear TM, Merranko J, Rosen D. How psychosocial well-being and usage amount predict inaccuracies in retrospective estimates of digital technology use. Mobile Media & Communication. 2020:2050157920902830.

33. Ofcom. Children and parents: media use and attitudes report. 2017.

34. Booker CL, Skew AJ, Kelly YJ, Sacker A. Media Use, Sports Participation, and Well-Being in Adolescence: Cross-Sectional Findings From the UK Household Longitudinal Study. Am J Public Health. 2014(0):e1–e7.

